# Profiles of the composite biomarker specific for β-amyloid accumulation in the brain relevant to age and sex

**DOI:** 10.1101/2023.01.13.23284514

**Authors:** Kouji Satoh, Maremi Sato-Ueshima, Hiroyo Kagami-Katsuyama, Masakazu Nakamura, Akihiko Ogata, Mari Maeda-Yamamoto, Jun Nishihira

## Abstract

Accumulation of β-amyloid (Aβ) in the brain occurs in the early phase of Alzheimer’s disease (AD), without symptoms of cognitive decline. Therefore, early detection of the accumulation phase is essential to prevent or delay AD. In this study, we investigated the effects of age, sex, apoprotein-E (ApoE) genotype and cognitive dysfunction on the ratio of Aβ_1-42_, Aβ_1-40_, and amyloid β precursor protein (APP)_669-711_ (composite biomarker: CM) in plasma using a sensitive Time of flight mass spectrometry (TOF-MS). In healthy subjects, the average CM value of males aged 30– 59 years was significantly higher than that of females, but no difference was observed between those aged 60 and 79 years. CM values increased considerably after 50 years of age, especially in females. The effect of the ApoE4 genotype on CM was greater in females than in males. The CM value of patients with AD was higher than that of healthy subjects and that of patients with Parkinson’s disease (PD). The CM value of patients with PD complicated by AD was higher than that of patients with PD. These results suggest that the CM value is influenced by age, sex, ApoE4 genotype, and central nervous system disorder. In addition, the CM value could be a reliable biomarker to distinguish not only patients with AD and healthy subjects but also AD and patients with PD. Since the measurement of CM is less invasive to the subjects, the method is helpful for the early detection of accumulation of Aβ before the appearance of AD symptoms. Moreover, this practical method can be used for differential diagnosis among a variety of patients with dementia.

## Introduction

The World Health Organization (WHO) reports that dementia affects more than 55 million people worldwide, and 60–70% of these people have Alzheimer’s disease (AD). In addition, treatment for AD is crucial because AD can also occur in other central nervous system disorders [1-3]. In the brain, an early symptom of AD is the accumulation of amyloid β (Aβ), produced by mutations in β- and δ-secretase [2,4,5]. Aβ accumulation triggers pathological findings in AD, including amyloid plaques and neurofibrillary tangles (NFTs). NFTs interact abnormally with cellular proteins and inhibit neuronal function. The hyperphosphorylation of tau occurs downstream of Aβ accumulation and activates Aβ synthesis. In the late phase of the neurodegenerative cascade in the AD brain, cholinergic deficits occur due to choline acetyltransferase deficiency. In addition, excitotoxicity, defined as an overexposure to glutamate or overstimulation of the N-methyl-D-aspartate (NMDA) receptor, plays an essential role in progressive neuronal loss in AD[2]. In this way, AD causes the accumulation of Aβ in the early stages, which leads to a cognitive decline in the brain.

Various factors influence AD development, including sex differences and genotypes of the ApoE gene [5,6]. Sex differences are one of the factors that affect the development of AD, and their effects are broad and include brain structure, stress response, and sex hormones. Serum and brain concentrations of the male hormone testosterone were significantly lower in patients compared to healthy individuals. In addition, endogenous testosterone deficiency caused by orchiectomy causes an increase in soluble Aβ concentrations in rat brains, and androgen administration reduces Aβ concentrations [5,7]. The female hormone, estrogen, is also involved in the risk of developing AD. Decreased estrogen levels in adulthood increase the risk of developing AD. In addition, ovariectomized mice show elevated levels of soluble Aβ in the brain, followed by an accelerated accumulation of Aβ with rapid worsening of cognitive behavior.[5,7].

With regard to the ApoE gene, people with one copy of E4-allyl in the ApoE gene have a nearly three-fold higher risk of developing AD, and those with two copies have an 8-12 times higher risk than those without the copy [5]. Furthermore, with regard to sex differences, females with E4-allyl have faster cognitive decline than males [5,8].

Although there are many patients with AD worldwide, effective treatments and therapeutic drugs are unavailable. During the discovery of therapeutic drugs, drugs for patients with early signs of cognitive declines, such as minor cognitive decline, have been anticipated. However, to date, only a few effective drugs are available. Therefore, the development of therapeutic drugs to be used before cognitive decline (preclinical phase) is currently in progress[2]. The accumulation of Aβ and formation of NFTs in the brain occur before signs of cognitive decline. In this context, measuring the accumulation of Aβ in the brain using central spinal fluid (CSF) or positron emission tomography (PET) is critical. However, these specialized techniques are unfavorable because of their high cost, invasiveness, and limited availability in routine clinical practice.

In 2018, Nakamura et al. measured Aβ concentration in plasma using mass spectrometry and showed 90% diagnostic accuracy for AD [9]. They demonstrated that Aβ_1-42_ concentrations decreased in the plasma of patients with AD, while Aβ_1-40_ and APP_669-711_ concentrations remained unchanged. Furthermore, the ratio of (Aβ_1-40_/ Aβ_1-42_) to (APP_669-711_ / Aβ_1-42_) (composite biomarker value: CM value) correlated with the accumulation of Aβ in the brain. By measuring Aβ in plasma, it has become possible to manage many samples in a limited period without invasiveness.

In this study, we measured the CM of patients (n = 112) with central nervous system disorders and healthy individuals (n=603). In addition, we investigated the change in CM value with age, sex, ApoE, and between patients with AD and healthy subjects. Here, we show that CM is a valuable biomarker for the definition of AD in healthy individuals and patients with AD and Parkinson’s disease (PD) patients.

## Materials and Methods

### Subjects

The subjects were essentially healthy registered volunteers managed by the Health Information Research Center at Hokkaido Information University. They thoroughly understood the significance, contents, and purposes of this study, “Accumulation and analysis of biomarkers involved in the prevention and improvement of dementia in healthy subjects,” and provided written consent. Healthy Japanese adult males and females between 30 and 79 years (n = 603) participated in the study. The data included age, sex, family history of dementia, blood chemistry results, and other clinical data. On the other hand, Japanese patients with central nervous system disorders between the ages of 50 and 91 (n = 112) joined the study after consultations with registered neurologists at the Hokkaido Neurosurgical Memorial Hospital (Sapporo, Japan). Written consent was obtained from patients or family members to participate in further studies.

### Laboratory tests

Blood samples were collected from healthy subjects and patients and sent to the Sapporo Clinical Laboratory, Inc. (Sapporo, Hokkaido, Japan). Complete blood count, liver function (aspartate aminotransferase, alanine aminotransferase, gamma-glutamyl transpeptidase, alkaline phosphatase, and lactate dehydrogenase), kidney function (blood urea nitrogen, creatinine, and uric acid), lipid metabolism (total cholesterol, low-density lipoprotein, high-density lipoprotein, and triglyceride), and glucose metabolism (blood glucose and hemoglobin A1c) was performed. We also measured the fatty acids (dihomo-γ-linoleic acid (DHLA), arachidonic acid (AA), eicosapentaenoic acid (EPA), docosahexaenoic acid (DHA), Zn, and 25-OH vitamin D.

### Aβ measurement

Peripheral blood Aβ levels were measured according to the method of Nakamura et al. [9]. Whole blood was collected in 7 mL EDTA-2Na tubes (Venoject II, TERUMO, Shibuya, Tokyo, Japan) and centrifuged at 2400 g for 5 min at room temperature within 60 min after blood collection. Plasma (500 μL) was transferred to storage tubes (96 Jacket Tubes 0.7 mL, FCR&Bio, Kobe, Hyogo, Japan) and frozen in a -80°C freezer. After they were isolated and enriched from abundant plasma proteins by immunoprecipitation, the specific affinity of the antibody plasma Aβ levels was measured using IP–MS (immunoprecipitation-mass spectrometry, Shimadzu Corporation, Kyoto, Kyoto, Japan). They used an analytical technique to quantify Aβ-related peptides of different masses using MALDI-TOF mass spectrometry (matrix assisted laser desorption/ionization-time of flight mass spectrometry, Shimadzu Corporation, Kyoto, Kyoto, Japan). Shimadzu Corporation blindly performed IP–MS analysis.

### Genotyping of ApoE gene

Whole blood was collected in a 4 mL EDTA-2K tube (Venoject II, TERUMO), and DNA was isolated from 400 uL whole blood using a QIAamp DNA Blood Minikit (QIAGEN, Hulsterweg, Venlo, Netherlands). We quantified the DNA content in the concentrated solution using Nano-Drop (Thermo Fisher, MA, USA). The genotype of the ApoE gene was analyzed by the Probe PCR method using two TaqMan probe sets (C_60538594A_10, C_ 60538594 B _20, Thermo Fisher, MA, USA). The reaction solution was mixed with 1ng/uL DNA solution. ApoE was analyzed using predesigned TaqMan genotyping assays (Applied Biosystems, CA, USA). Approximately 1 ng of genomic DNA was amplified in a 5 μL reaction mixture in a 96-well plate containing 1 × Probe PCR ExTaq master mix (Takara Bio, Shiga, Japan) and 1 × Taqman probe mix containing the respective primers and probes. QuantStudio 3 was used with a standard endpoint genotyping program (Thermo Fisher Scientific, MA, USA). According to the manufacturer’s protocol, PCR was performed with an initial denaturation at 94°C for 5 min, followed by 40 two-step cycles (denaturation for 30 s at 94°C, primer annealing, and extension for 60 s at 60°C).

### Statistical analysis

All statistical analyses were performed using the Statistical Package for Social Science (SPSS® Statistics version 25, IBM Corp, NY, USA). Statistical significance was set at p < 0.05. The age, sex, and ApoE genotype of healthy subjects were randomly divided into several groups. We used the Mann–Whitney U test or Kruskal– Wallis H test to compare two or more multiple groups.

### Ethics

This study was conducted in accordance with the guidelines of the Declaration of Helsinki [10]. The ethics committee approved all procedures involving human subjects at Hokkaido Information University (for healthy people, approved on November 26, 2019; approval number: 2019-31; for patients, approved on July 31, 2020; approval number: 2020-19). Therefore, this study complied with the ethical guidelines for human medical research as per the Ministry of Education, Culture, Sports, Science, and Technology and the Ministry of Health, Labour, and Welfare.

## Results

### Effects of age and sex on composite biomarker values in healthy subjects

We investigated the CM values of healthy subjects between the ages of 30 and 79 years. The male-to-female ratio was approximately 2:5 (181 males, 422 females). The highest ratio was approximately 1:1 (male = 26, female = 29) between 70 and 79 years, and the lowest ratio was approximately 1:4 (male = 35, female = 135) between 50–59 years. Most subjects were aged between 50 and 59 years, and the smallest group was aged between 70 and 79 years (Table 1). Healthy subjects were divided into two groups, male and female, to analyze the effects of age and sex on the CM values. We subdivided these two groups into five age groups (30–39 years, 40–49 years, 50–59 years, 60–69 years, and 70–79 years of age) (10 groups in total) and then calculated the average CM of each group. For the group-to-group comparison, we used the Mann–Whitney U test (Table 1, Fig 1). Under 59 years of age, the CM value of males was higher than that of females (30–39 years: p = 0.038, 40–49 years: p = 0.013, 50–59 years: p = 0.007). However, the CM value of males over 60 years of age was similar to that of females (Table 1). The CM value increased with age, especially after 50 years of age (males: p = 0.038, comparison between 40–49 and 50–59, females: p = 0.006 and 0.018 comparisons between 40–49 and 50–59, between 50–59 and 60–69, Fig 1). The results showed that the standard deviation of the CM values increased for both males and females aged 60 years and older, especially in females (Table 1).

**Table 1.** The effect of CM value on sex and age.

| Age | Sex | No. of Subjects | Ave. of CM | Std |
| --- | --- | --- | --- | --- |
| 30-39 | male | 20 | -0.547 | 0.349 |
|  | female | 53 | -0.741 | 0.391 |
| 40-49 | male | 37 | -0.509 | 0.341 |
|  | female | 102 | -0.694 | 0.333 |
| 50-59 | male | 35 | -0.357 | 0.382 |
|  | female | 135 | -0.535 | 0.429 |
| 60-69 | male | 63 | -0.276 | 0.474 |
|  | female | 103 | -0.335 | 0.623 |
| 70-79 | male | 26 | -0.076 | 0.644 |
|  | female | 29 | -0.097 | 0.718 |

**Fig 1.**
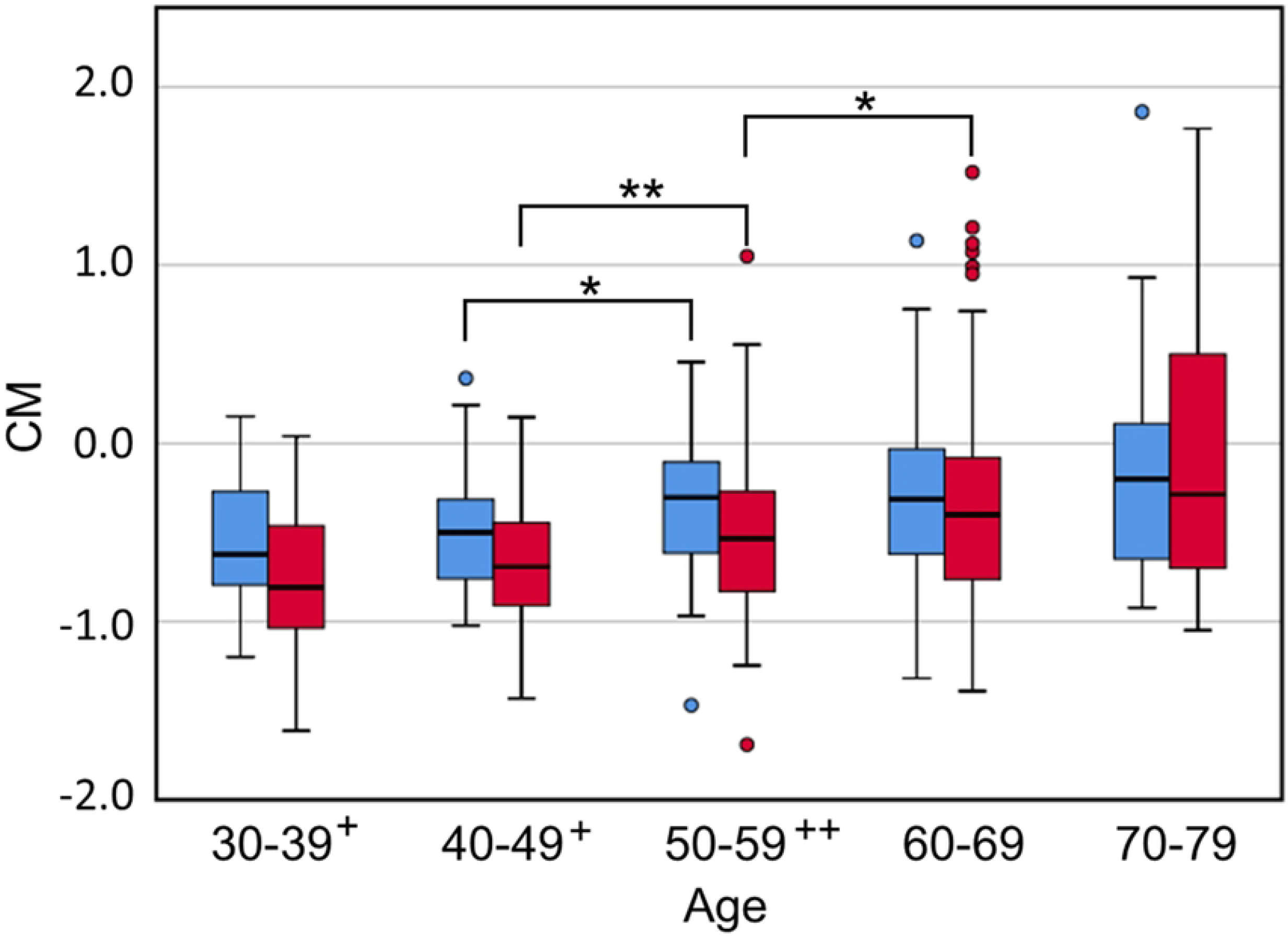
The relationship of CM value on sex and age. The bar plot showed the distribution of the CM values of the healthy subjects. The number of subjects is listed in Table 1. The difference in the distribution of CM values between males (blue) and females (red) was examined using the U-test (+: p ≤ 0.05, ++: p ≤ 0.01). The difference in the CM value distribution between ages of the same sexgender was also examined using the U-test (*: p ≤ 0.05, **: p ≤ 0.01). X-axis: age. Y-axis: CM value.

We surveyed subjects between 60 and 79 years of age to determine the relationship between the CM value and the subjects’ ages in males and females (Fig 2). The frequency of subjects with CM ≥ 0 was similar between male and female (male = 26/89 (29.2%), female = 32/132 (24.2%)). However, in subjects with CM ≥ 0, the frequency of subjects with CM > 0.5 was significantly higher in females than in males(male = 9/26 (34.6%), female = 20/32 (62.5%), p = 0.035, χ^2^test). This result suggests that the females are more dominant aged people with higher CM values.

**Fig 2.**
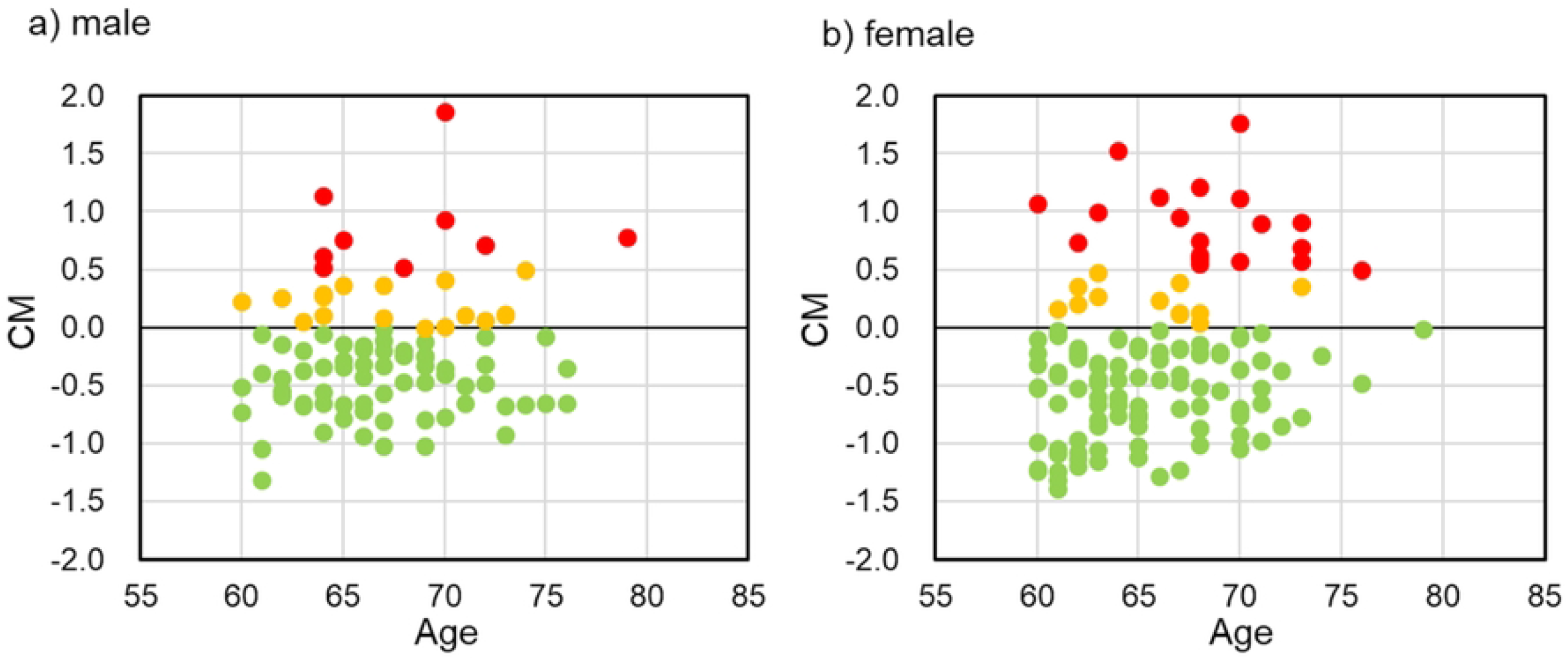
The distribution of CM value of subjects ages 60 to 79 years. It shows the distribution of CM values among healthy subjects aged 60 to 79 years. a: Male (n = 89);, b: Female (n = 132). Green: CM value < 0; yellow: 0 ≤ CM value ≤ 0.5; red: CM value > 0.5. X-axis: age; Y-axis: CM value.

### Influence of genotypes on composite biomarker values in healthy subjects

We investigated the relationship between ApoE4 genotypes and CM values of healthy subjects. Healthy male subjects tended to have higher CM values in the population with ApoE4; however, there was no significant difference between subjects with ApoE4 and those without ApoE4 (Fig 3). Among healthy female subjects, CM values were higher in the ApoE4 group (p = 0.078, U-test) from 50–59 years of age. There was a significant difference between the group with ApoE4 and those aged 60 years or older without ApoE4 (p = 0.026, U-test) (Fig 3).

**Fig 3.**
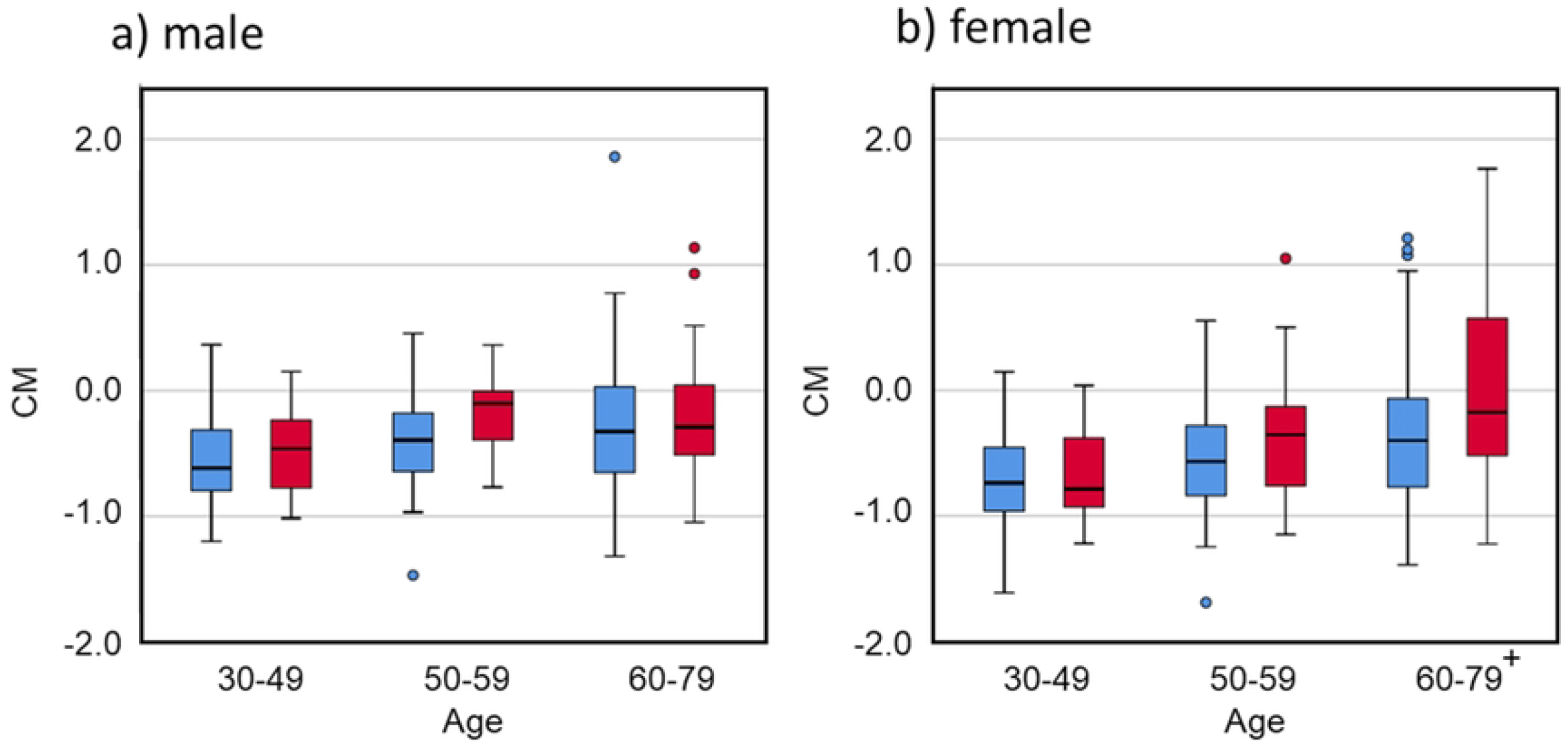
The relationship between CM value and ApoE genotype. The bar plot shows the distribution of the CM values of the healthy subjects. In addition, the difference in the distribution of CM values between subjects with E4 allyl (red) and those without E4 allyl (blue) was analyzed using the U-test. a) The number of male subjects was 52 for those aged 30–49 years (E4 allyl: 15, no E4 allyl: 37), 35 for those aged 50–59 years (E4 allyl: 8, no E4 allyl: 27), and 87 for those aged 60–79 years (E4 allyl: 23, no E4 allyl: 64). b) The number of female subjects was 151 for those aged 30– 49 years (E4 allyl: 31, no E4 allyl: 120), 133 for those aged 50–59 years (E4 allyl: 25, no E4 allyl: 108), and 128 for those aged 60–79 years (E4 allyl: 21, no E4 allyl: 107). +: p < 0.05. X-axis: 30–49: 30–49 years old, 50-59: 50–59 years old, 60–79: 60–79 years old. Y-axis: CM value.

### Comparison of CM values between healthy subjects and patients with central nervous system disorder

We compared the CM values of healthy subjects aged 60–79 years (n = 221, male = 89, female = 132, average age = 66.4) and patients with central nervous system disorders (n = 112, male = 49, female = 63, average age = 77.9), includingAD, argyrophilic grain dementia, corticobasal degeneration, dementia with Lewy bodies, mild cognitive impairment, multiple system atrophy, normal pressure hydrocephalus, Parkinson’s disease (PD), Parkinson disease with dementia (PDD), progressive supranuclear palsy, senile dementia, vascular dementia, and other diseases with dementia (Table 2). In the ages of 60 to 79 years, the CM value of patients was higher than that of healthy subjects (patients: CM = 0.501, healthy: CM = -0.257, p = 0.000). Furthermore, from the distribution of CM values, 75% or more of healthy subjects between the ages of 60 and 79 years had a CM value of less than 0, whereas more than half of the patients had a CM value of 0 or more (Fig 4).

**Table 2.** The number of healthy subjects and patients with central nervous system disorders.

|  | Total | male | female |
| --- | --- | --- | --- |
| Healthy | 221 | 89 | 132 |
| Patients | 112 | 49 | 63 |
| AD | 13 | 2 | 11 |
| PD | 33 | 14 | 19 |
| PDD | 23 | 12 | 11 |
| PD+AD | 18 | 7 | 11 |
| others | 25 | 14 | 11 |

AD: Alzheimer's disease, PD: Parkinson's disease, PDD: PD with dementia,
PD+AD: PD with AD.

**Fig 4.**
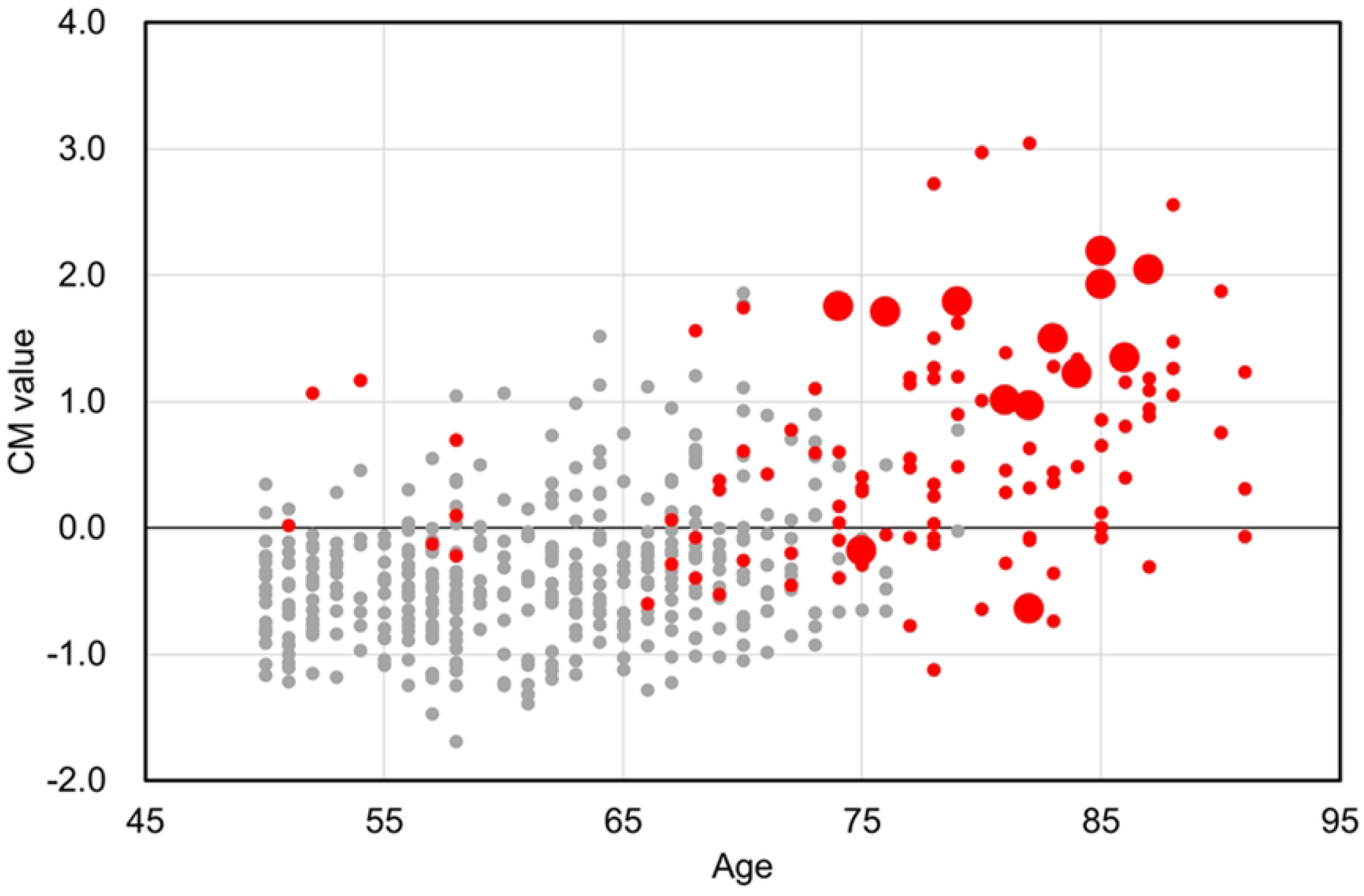
The distribution of CM value in healthy subjects and patients with dementia. The distribution of CM values of healthy subjects (50–79: gray) and patients with central nervous system disorders (red). The large red circles indicate the CM values of patients with AD. X-axis: age; Y-axis: CM value.

We compared the CM values among healthy subjects, patients with AD (n = 13, male = 2, female = 11), patients with PD (n = 33, male = 14, female = 19), patients with PDD (n = 23, male = 12, female = 11), and patients with PD+AD (n = 18, male = 7, female = 11) using the Kruskal Wallis H test and Mann-Whitney U test (Table2). The CM values among the healthy, AD, and PD groups differed significantly (p = 0.000). The average CM value in healthy subjects was -0.257, and those in patients with AD, PD, PDD, and PD+AD were 1.281, 0.406, 0.512, and 0.720, respectively (Fig 5). The CM value in AD patients was significantly highest, and the value in healthy subjects was significantly lowest. The CM values with PDD patients were similar to those in PD patients (p = 0.344), but those with PD+AD patients were significantly higher than those in PD patients (p = 0.043).

**Fig 5.**
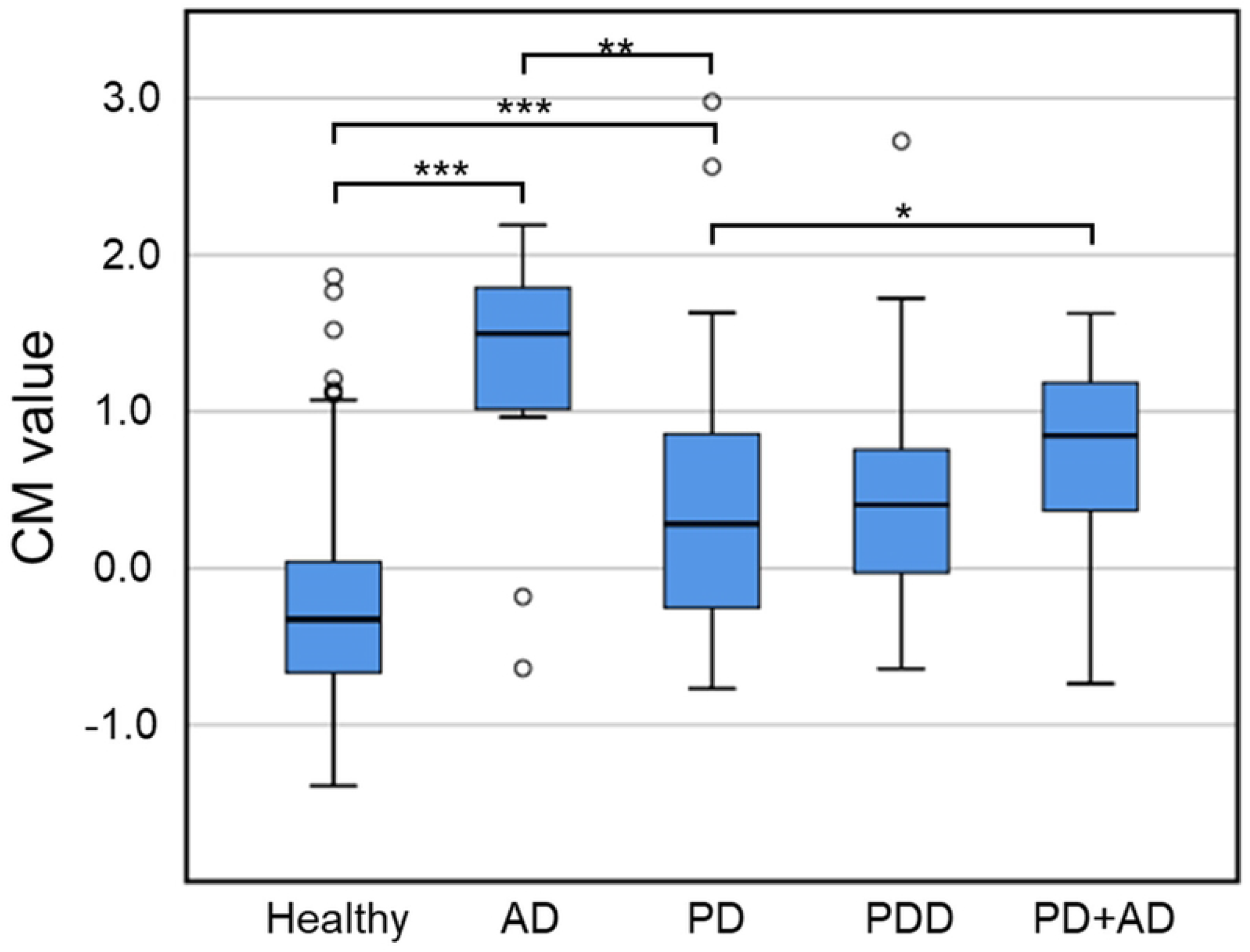
The distribution of CM value in each disease group. The bar plot shows the distribution of CM values of healthy subjects (n = 221), patients with AD (n = 13), PD (n = 33), PD with dementia (PDD, n = 23), and PD+AD (n=18). The number of participants is shown in Table 2. The U-test calculated the difference in the CM values between each other. *: p < 0.05, **: p < 0.01, ***: p < 0.001. Y-axis: CM value.

## Discussions

In 2019, Nakamura et al. measured APP_669-711_, Aβ_1-40_, and Aβ_1-42_ in peripheral blood and calculated the CM value from the ratio of their concentrations. Since the CM value is 0.376 or higher and contains approximately 90% of patients with AD [9], detecting the accumulation of Aβ in the brain with high sensitivity has become possible.

Furthermore, this method uses plasma from available blood collection, with a minimal burden on the patient [9]. In this study, we carried out CM measurements for a large number of people, including not only patients with dementia but also healthy subjects, and the correlation between CM value and age, sex, and central nervous system disorders (especially AD and PD) was analyzed.

### The influence of age, sex, and genotype on CM value

In this study, we first measured the CM values of healthy subjects. The CM levels tended to increase with age. In subjects younger than 60 years, the male group had significantly higher CM values than the female group; however, there was no difference in CM values between males and females over 60 years (Fig 1). In males, the CM value increased consistently with age; however, in females, the CM value increased after 50 years of age. These results suggest that after the age of 50 years, changes in blood Aβ levels differ between males and females.

One of the factors contributing to these sex differences may be sex hormone levels. In males, blood testosterone concentration and the risk of developing AD have been correlated, based on a report that male patients with AD have lower serum testosterone levels [11,12]. In addition, an inverse correlation exists between testosterone and Aβ concentrations in the brain [13]. These findings suggest that a decrease in testosterone levels promotes the accumulation of Aβ in the brain [6,7,11-13]. In addition, testosterone levels decrease linearly with the age of 20 in males [14,15]. These findings imply that the accumulation of Aβ in the brain increases consistently with age, and the consistent increase in the brain is similar to the change in the CM value of males. In females, reduced estrogen levels appear to be associated with an increased risk of AD [6,16]. Estrogen levels decrease sharply during menopause [14]. These facts indicate that the accumulation of Aβ in the brain increases sharply after 50 years of age, which is similar to the change in the CM value of females. Moreover, at the age of 60-79, the proportion of male subjects with high CM values was higher than that of female subjects (Fig 2). The standard distribution of CM values in females was greater than that in males (Table 1). These facts suggest that the CM value in female subjects may be influenced by sex hormones, as well as other factors.

In this study, we found a significant increase in the CM value due to the ApoE4 gene in females aged 60-79 (Fig 3). This suggests that the effect of ApoE4 on the CM value was different between males and females. Individuals with the ApoE4 gene have an increased risk of developing AD [5], and the influence of the ApoE4 gene is present in both males and females. However, the effect of ApoE4 on the onset of AD may differ between males and females [5]. In addition, the effects of ApoE4 were more pronounced after menopause in females. This suggests that the interaction of estrogen with ApoE4 increases the risk of developing AD [5]. These reports imply that the accumulation of Aβ in the brain is more pronounced in females carrying the ApoE4 gene. This enhancement of Aβ accumulation in women by the ApoE4 gene is similar to the increase in CM values in females with the ApoE4 gene. This similarity between the change in CM value in blood and the accumulation of Aβ in the brain indicates that, in healthy people, the CM value in the blood reflects the accumulation of Aβ in the brain.

### The difference in CM value between healthy subjects and patients with central nervous system disorder

We measured the CM values in 112 patients with central nervous system disorders. Certified neurologists diagnosed patients with central nervous system disorders (Table 2). The CM values of healthy subjects were lower than those of patients with central nervous system disorders. Therefore, we can distinguish CM values between healthy individuals and patients with central nervous system disorders. Moreover, the average CM value in patients with AD (1.282) was significantly higher not only in healthy subjects (−0.257) but also in patients with PD (0.406). Notably, the CM value in patients with PD was higher than that in healthy subjects. This result implies that the CM value can be distinguished between healthy individuals, patients with central nervous system disorders, and patients with either AD or PD (Fig. 5). In addition, this result implies that patients with a central nervous system disorder, regardless of the type of disease, may have a more accumulation of Aβ in the brain than healthy individuals. PD and dementia with Lewy bodies are categorized into a group of diseases called α-synucleinopathy, in which toxic oligomeric α-synuclein accumulates in nerve cells. In patients with PD, the concentration of α-synuclein in the CSF is lower than that in healthy individuals [17]. It is positively correlated with the concentration of Aβ_1-42_ in the CSF [18]. These reports imply that the concentration of Aβ_1-42_ in the blood is lower in patients with PD than in healthy individuals. Therefore, patients with PD may have higher CM values than healthy individuals. We also found that the CM value in patients with PD complicated by AD (0.720) was significantly higher than in patients with PD (Fig 5). This result suggests that the CM value measured by this method can be used for early detection of AD in healthy subjects and differential diagnosis of patients complicated with other central nervous system disorders. Thus, the CM value is a valuable biomarker for detecting AD and other central nervous system disorders, with a decrease of Aβ_1-42_ in CSF.

## Conclusion

We demonstrated that the CM value calculated from Aβ concentration in the blood is an index for distinguishing between patients with central nervous system disorders and healthy individuals. Moreover, the CM value differentiates patients with AD from patients with PD and healthy people. As the measurement of Aβ in the peripheral blood used in this study has a significant advantage with little burden on subjects, it is possible to measure patients with AD and healthy people. Furthermore, measurements over time can be used to investigate changes in the speed of Aβ accumulation, which helps determine the preclinical phase.

## Data Availability

Data Availability Statement: The data obtained from the “Sukoyaka Health Survey” is not cur-rently in a publicly accessible repository, but is scheduled to be released in 2024 from a publicly accessible repository managed by the DDBJ (DNA Data Bank of Japan) JGA (Japanese Geno-type-phenotype Archive https://www.ddbj.nig.ac.jp/jga/index-e.html).

## Acknowledgments

We thank members of Hokkaido Information University, Center of Health Information Science, and Hokkaido Neurosurgical Memorial Hospital for their technical assistance with the clinical trial and data treatment. This study was supported by the cross-ministerial Strategic Innovation Promotion Program (SIP), “Technologies for Smart Bio-industry and Agriculture” and Public/Private R&D Investment Strategic Expansion PrograM (PRISM).

